# Cross-sectional and longitudinal associations between dietary intake and depressive symptoms in young South African adults: The African-PREDICT study

**DOI:** 10.64898/2026.02.13.26346223

**Authors:** Esmé Jansen van Vuren, Adrienne O’Neil, Deborah N Ashtree, Melissa M Lane, Rebecca Orr, Marlien Pieters, Tertia van Zyl

## Abstract

**Introduction:** Depression is highly prevalent among young adults worldwide. While research links health behaviours, such as dietary intake, to depression, few studies have examined these associations among young adults in low- and middle-income countries, including South Africa. This study investigated whether dietary intake was associated with an increased risk of depression in a cohort of young South African adults, aged 20-30 years, as part of the Global burden of disease Lifestyle And mental Disorder (GLAD) project.

**Methods:** This five-year prospective cohort study was conducted in the North West Province of South Africa in accordance with the GLAD project protocol (DERR1-10.2196/65576). Dietary exposures were evaluated using three non-consecutive 24-hour dietary recalls, measuring daily intake of various food groups and nutrients as defined by the Global Burden of Disease study. Depression outcomes were assessed at baseline (N=1039) and follow-up (N=551) using the Patient Health Questionnaire (PHQ-9, cut-off ≥10). Logistic and Poisson regression analyses were performed, with results presented as odds ratios (OR) and relative risk ratios (RR), respectively. Four models were run: unadjusted, sociodemographic-adjusted, total energy (TE) intake-adjusted and fully adjusted (including sociodemographic information and TE intake). For longitudinal analyses of incident depression, baseline depression cases were additionally excluded (n=403).

**Results:** Participants (average age 24.55 years) had a balanced distribution of sex (51.4% female) and race (48.6% Black), and a 29.45% baseline prevalence of depression. Higher milk intake was associated with a lower risk of incident depression (RR=0.94, 95% CI 0.91-0.98) in the TE-adjusted longitudinal model. Cross-sectionally, higher sugar-sweetened beverage consumption associated with higher odds of depression, while higher calcium intake (OR=0.48, 95% CI 0.31; 0.76) and vegetable consumption (OR=0.74, 95% CI 0.61, 0.91) were associated with lower odds of depression after TE intake adjustment. Higher fibre intake was associated with lower odds of depression in the unadjusted model.

**Conclusion:** Higher daily milk intake was associated with a lower risk of depression, while higher calcium, vegetable, and fibre intake were associated with a lower prevalence of depression in young adults. These findings suggest that prevention strategies for common mental disorders could include dietary approaches within mental health care.

## 1. Introduction

The global burden of mental disorders is substantial. In 2021, common mental disorders, namely depressive and anxiety disorders, were ranked among the leading causes of disability-adjusted life years (DALYs) and were major contributors to years lived with disability [1]. Major depressive disorder (MDD) remains a significant contributor to this burden, accounting for the highest proportional age-standardized incidence rate (approximately 76%) of all mental disorder subtypes in 2021 [1]. Mental disorder-related DALYs peak between the ages of 25 and 34 years, highlighting the heightened vulnerability of young adults [2]. Life course data further demonstrate that the onset of depression and anxiety often occurs at younger ages, and that age-specific interventions targeting modifiable risk factors are needed to address the global mental health burden [3].

Given the substantial burden of common mental disorders, dietary intake is a relevant modifiable exposure and is already incorporated within the Global Burden of Disease (GBD) study as a risk factor for non-communicable diseases, including cardiovascular disease (CVD), which commonly co-occurs with depression and anxiety [4]. Emerging evidence also suggests that diet may serve as a modifiable risk factor for common mental disorders, such as depression and anxiety [6, 7]. However, lifestyle exposures are not yet routinely reported as risk-outcome pairs for these common mental health conditions within the GBD framework, limiting evidence-informed prevention priorities and guideline development, particularly in low- and middle-income countries (LMICs). This gap provides the rationale for the Global burden of disease Lifestyle And mental Disorder (GLAD) Taskforce, an international collaborative initiative aiming to integrate lifestyle exposures as risk factors for common mental disorders in the GBD study [8].

A key objective of the GLAD project is to produce evidence on the diet-common mental disorder association in regions of the world where data are particularly scarce, including LMICs, such as South Africa. This evidence gap is concerning given the rising prevalence of these depression in LMICs [2, 9]. In 2021, sub-Saharan Africa had one of the highest age-standardized incidence rates for mental disorders, with Southern sub-Saharan Africa reporting the fourth highest proportion of DALYs due to MDD [1]. These trends, alongside a significant dietary transition in Black South Africans from prudent dietary patterns, high in minimally processed plant-based foods and moderate animal-based foods, toward more Westernized diets high in refined sugars, saturated fats, and ultra-processed food (UPF) [10], underscore the urgent need for context-specific research. Accordingly, this study aims to investigate whether cross-sectional and longitudinal associations exist between depression and the consumption of specific food groups and nutrients in a cohort of young South African adults.

## 2. Methods and materials

### 2.1. Study design

This study formed part of the GLAD Taskforce [8] and used data from the African Prospective study on the Early Detection and Identification of Cardiovascular disease and Hypertension (African-PREDICT). The African-PREDICT study was initiated to monitor the development of hypertension in a bi-racial sample residing in the JB Marks local municipality, North West province, South Africa over a five-year follow-up period [11]. The study’s baseline phase was conducted from 2013 to 2017 and included 1202 young individuals aged 20-30 years. Exclusion criteria included clinic brachial blood pressure >140/90mmHg; HIV infection; previous diagnosis of any of the following chronic diseases: cancer, tuberculosis, liver disease, renal disease, diabetes or CVD; medication use for hypertension, diabetes, or HIV; presence of fever; being pregnant or breastfeeding; phobia of needles; inability to read or understand English; and any races other than self-reported Black or White racial groups. The study’s first follow-up phase commenced in 2018 and was completed in 2024, during which each participant completed a single follow-up assessment.

For the present analyses, individuals with missing dietary (n=31) or depressive symptoms (n=132) data at baseline were excluded, resulting in a baseline analytical sample of 1039 individuals. Of these individuals, 710 were successfully followed up, after which individuals with missing depressive symptom data at follow-up (n=159), resulting in a final longitudinal sample of 551 individuals as shown in Figure 1. For incident depression analyses, those who met depression criteria at baseline (see description below) were further excluded (n=148).

**Figure 1.**
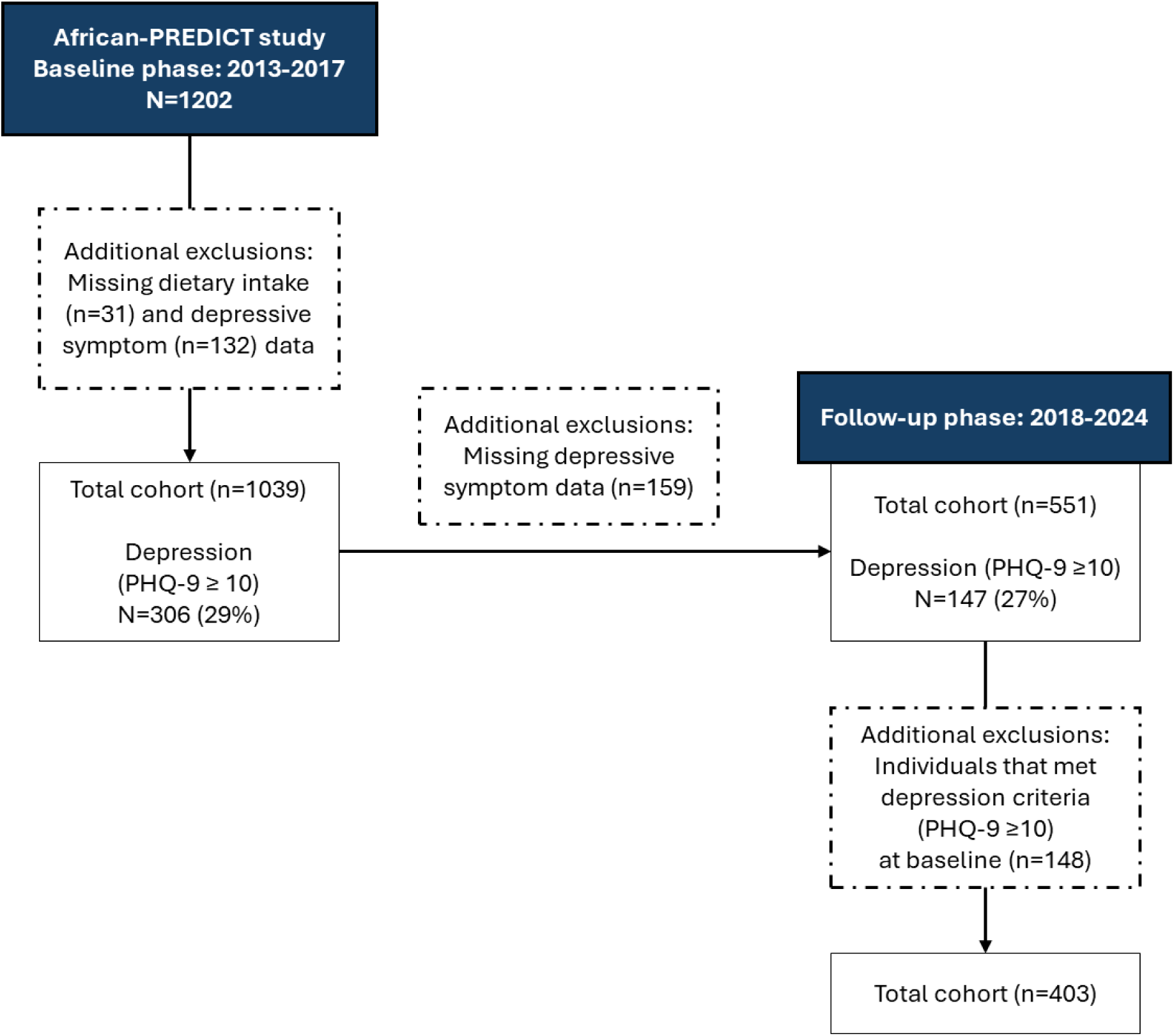
Study design flow diagram.

The African-PREDICT study complies with the Declaration of Helsinki, was approved by the Health Research Ethics Committee of the North-West University (NWU-00001-12-A1) and is registered on Clinical-Trials.gov (NCT03292094). Written informed consent was provided by all participants prior to data collection.

### 2.2. Organizational procedures

Screening procedures were conducted to determine eligibility for participation according to the aforementioned inclusion and exclusion criteria at baseline. Eligible participants were invited to the Hypertension Research and Training Clinic, situated on the Potchefstroom campus of North-West University, for baseline and follow-up data collection. For both phases, participants were requested to fast from 22:00pm on the evening prior to their arrival at the clinic. A maximum of four participants arrived at the clinic at approximately 08:00 on the day of participation, after which all procedures and measurements were reiterated. Participants had the opportunity to ask questions, after which they provided written informed consent. A registered research nurse supervised all procedures and collected fasting blood samples early in the morning. Various measurements and assessments were completed throughout the day (described below). The participants also received direct feedback on selected measurements and if any health-related irregularities were identified, participants were referred to a healthcare provider.

### 2.3. Depressive symptom assessment

Depressive symptoms were assessed at baseline and follow-up using the validated nine-item Patient Health Questionnaire (PHQ-9), which is based on the diagnostic criteria for MDD outlined in the Diagnostic and Statistical Manual of Mental Disorders, Fourth Edition (DSM-IV), and retained in the current Fifth Edition (DSM-5-TR) [12]. Each item is rated on a 4-point Likert scale (0 = “not at all” to 3 = “nearly every day”), yielding a total score ranging from 0 to 27, with higher scores indicating more severe depressive symptoms. Internal consistency was evaluated using the nine items using Cronbach’s alpha (α) and McDonald’s omega (ω). The scale has demonstrated good reliability (α = 0.82; ω = 0.82). A cut-off score of ≥10 was used to indicate moderate-to-severe depressive symptoms [13], a threshold shown to correlate well with a clinical diagnosis of MDD across adult age groups [14, 15].

### 2.4. Dietary data

Three 24-hour dietary recall interviews were conducted at baseline by trained fieldworkers. The first interview was conducted on the same day as the other study measurements, and the two follow-up interviews were conducted on two separate days within the following week, including one weekend day to obtain information on weekend food consumption. A standardized dietary collection kit was used, containing visual aids such as pictures, food packages, household measurement tools, and food models. Food portion sizes were estimated using plastic food models, household utensils, food packaging materials, and a portion-size photograph book. The South African Medical Research Council (SAMRC) food composition tables [16] were used to code the dietary recalls, and the food quantities manual [17] was employed to convert household measures into grams. When specific food items were unavailable in the tables, they were purchased, weighed, and documented for future reference. Final codes and quantities for each dietary record were cross-checked against the original recall data to ensure accuracy in coding and data capturing. Nutrient analysis of the baseline dietary data was conducted by the SAMRC Biostatistics Unit using South African food composition tables [16]. The individual average intakes of energy and the GBD-defined dietary exposures were calculated from the three dietary recalls for each participant. These food groups and nutrients as defined by the GBD included fruit, vegetables, legumes, wholegrains, nuts and seeds, milk, red meat, processed meat, sugar-sweetened beverages, total fibre, calcium, and omega-3 fatty acids [eicosapentaenoic acid (EPA) and docosahexaenoic acid (DHA)] expressed in grams per day (g/day), and polyunsaturated fatty acids (PUFA) expressed as percentage (%) of total energy (TE) per day. Although the GBD study does not currently include data on UPF and non-UPF consumption, these variables were incorporated using the NOVA classification system and expressed separately as a % of TE intake [18]. To enhance the practical interpretability of the findings, the units of some of the food groups and nutrients were rescaled to reflect estimated daily consumption based on dietary intake data from South African adults [19–22]. Accordingly, fruit was expressed as increments of 100 g/day, vegetables as 100g/day, legumes as 25 g/day, wholegrains as 50 g/day, milk as 10 g/day, red meat as 25 g/day, processed meat as 17 g/day, sugar-sweetened beverages as 50 g/day, while all other nutrients were expressed as 1 g/day, PUFA and UPF each as % of TE.

### 2.5. Demographic and medical history questionnaires

A general health questionnaire was used to obtain sociodemographic information, including age, sex, race, total household income, highest level of education, and employment status. The Kuppuswamy’s Socioeconomic Status (SES) Scale [23] was adapted to calculate a composite socioeconomic score. Three scale comprises of three components, namely the total household income, highest level of education, and employment status (SASCO skill level). Participants received a total SES score ranging from 5 to 30 and were subsequently categorized into low (5-18 points), middle (19-24 points) or high (25-30 points) SES, as described previously [24].

### 2.6. Body composition

Body mass index (BMI) was calculated as body weight in kilograms (obtained with the SECA 813 Electronic Scale, SECA, Hamburg, Germany) divided by the square of body height in meters (obtained using the SECA 213 Portable Stadiometer, SECA, Hamburg, Germany).

### 2.7. Statistical analyses

Statistical analyses were performed using IBM® SPSS® Statistics version 30 (IBM Corporation; Armonk, New York, USA) and graphical figures were created using GraphPad Prism version 5.03 (GraphPad Software Inc., CA, USA). Descriptive statistics were computed, with continuous variables reported as means ± standard deviations (SD), and categorical variables as the counts (n) and percentages (%). Additionally, independent t-tests were used to compare participant characteristics and dietary exposures between individuals with and without depression at baseline.

Logistic regression analysis was performed to determine whether dietary consumption of specific food groups and nutrients was associated with increased odds of depression at baseline. Poisson regression analyses were then used to evaluate whether baseline dietary consumption was associated with the risk of incident depression at follow-up. For each regression analysis, four separate models were fitted as model 1) an unadjusted model, model 2) adjusted for sociodemographic covariates (age, sex, race, and SES), model 3) adjusted for TE intake using Willett’s residual method [25], and model 4) a fully adjusted model including both sociodemographic covariates and TE intake using Willett’s residual method. More than 90% of participants reported no consumption of legumes, nuts, or seeds. Consequently, these food groups were excluded from all regression analyses due to an insufficient number of valid cases. Sensitivity analyses were performed by additionally adjusting for body composition (BMI) in the final fully adjusted models (model 4). P-values were adjusted for multiple testing using the Simes procedure, with the number of hypotheses (m) set to 13 per model.

## 3. Results

### 3.1. Baseline Cohort Description

In the total cohort included at baseline (n=1039), the prevalence of depression was 29.45% (n=306). Participant characteristics are presented in Table 1 by depression status. The mean age of the participants was 24.55 years, with a balanced distribution of sex (male: 48.6%, female: 51.4%) and race (Black: 48.6%, White: 51.4%) in the total group. Race distribution differed by depression status, with a higher proportion of Black participants among those with depression (62.4%), and a higher proportion of White participants among those without depression (57.2%). Socio-economic status also differed by depression status, with 57,5% of individuals with depression were classified as low socio-economic status, whereas 36.7% of individuals without depression were classified as high socio-economic status. Overall, the average BMI was 24.13 kg/m^2^. No differences were shown between the groups for BMI. Individuals with depression had a lower TE intake compared to those without depression (mean difference: 579.11 kJ/day).

**Table 1.**
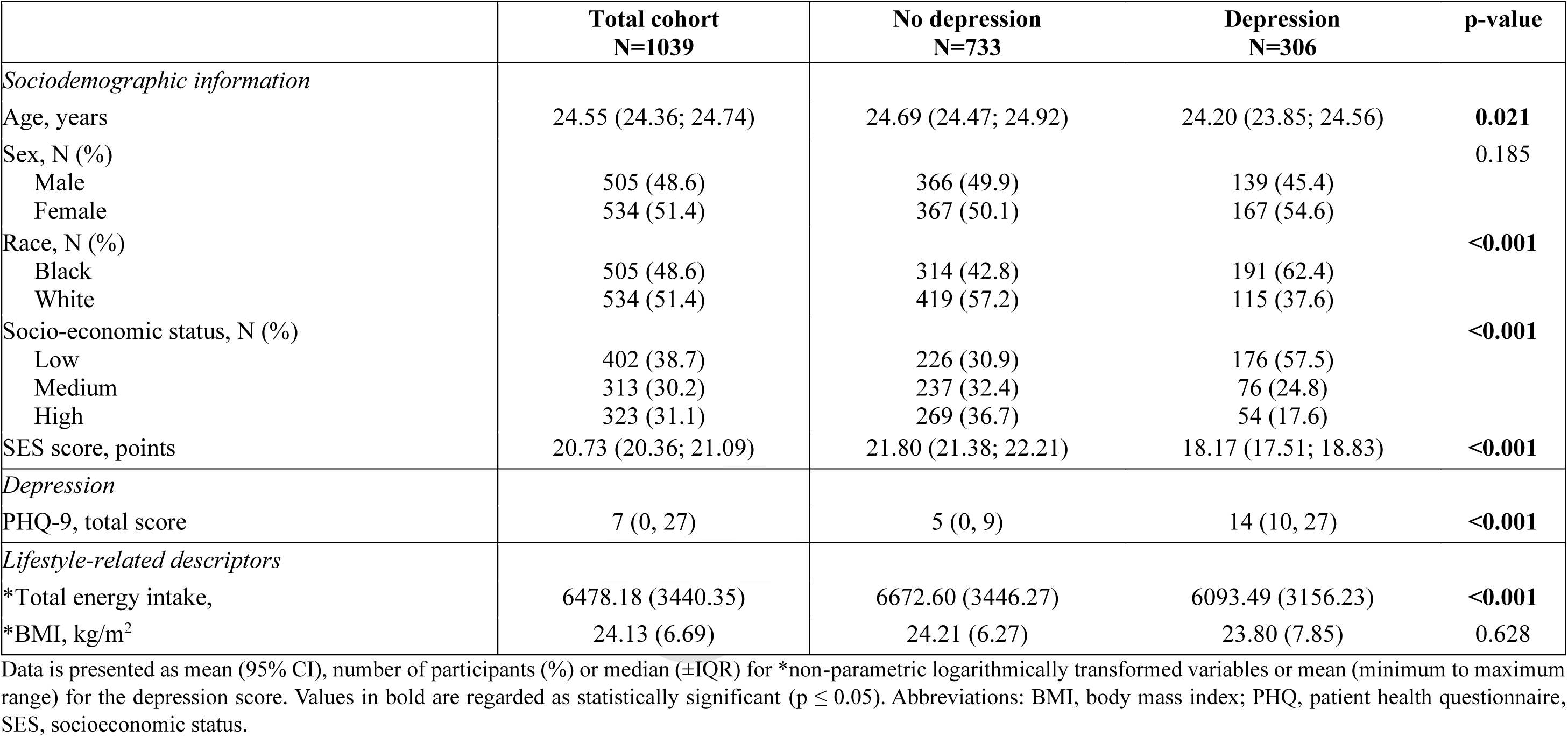
Baseline cohort description.

Table 2 presents the average intake of dietary risk factors at baseline. Individuals with depression had a lower intake of fruit (mean difference: 27.5 g/day), vegetables (mean difference: 25 g/day), legumes (mean difference: 13 g/day), wholegrains (mean difference: 12.5 g/day), fibre (mean difference: 0.64 g/day), PUFA (mean difference: 2.07 g/day) and calcium (mean difference: 0.12 g/day), as well as higher intake of sugar-sweetened beverages (mean difference: 33.34 g/day), when compared to individuals without depression (Table 2).

**Table 2.** Average intake of dietary risk factors at baseline.

|  | **Total cohort** | **No depression** | **Depression** | **p-value** |
| --- | --- | --- | --- | --- |
| Fruit intake, g/day (N=444) | 145.26 (150.85) | 100.00 (150.00) | 72.50 (116.67) | **≤0.001** |
| Vegetable intake, g/day (N=842) | 131.91 (114.54) | 113.33 (105.45) | 88.33 (109.68) | **≤0.001** |
| Legumes intake, g/day (N=102) | 61.94 (86.85) | 34.50 (63.25) | 21.50 (40.40) | **0.040** |
| Wholegrain intake, g/day (N=341) | 85.67 (85.71) | 62.50 (73.50) | 50.00 (52.29) | **0.016** |
| Nuts and seeds intake, g/day (N=61) | 30.29 (26.23) | 20.00 (38.33) | 23.33 (17.50) | 0.920 |
| Milk intake, g/day (N=719) | 168.44 (158.74) | 140.00 (181.75) | 110.00 (167.50) | 0.109 |
| Red meat intake, g/day (N=540) | 115.03 (96.39) | 100.00 (92.50) | 89.83 (92.92) | 0.205 |
| Processed meat intake, g/day (N=640) | 65.68 (60.78) | 46.67 (58.33) | 50.00 (63.33) | 0.404 |
| Sugar-sweetened beverages intake (N=641), g/day | 362.58 (385.94) | 333.33 (325.00) | 366.67 (358.33) | **0.025** |
| Fibre intake, g/day (N=1022) | 14.79 (8.00) | 13.27 (9.46) | 12.63 (7.74) | **0.001** |
| Calcium intake, g/day (N=1022) | 0.52 (0.36) | 0.48 (0.50) | 0.36 (0.38) | **≤0.001** |
| Omega-3 fatty acid intake, g/day (N=1022) | 168.77 (434.46) | 65.67 (126.83) | 53.50 (103.52) | 0.349 |
| Polyunsaturated fat intake, g/day (N=1014) | 17.03 (10.94) | 15.20 (13.19) | 13.13 (11.96) | **0.013** |
| Ultra-processed food intake, % of TE (N=1014) | 46.09 (18.15) | 45.09 (25.47) | 47.20 (23.94) | 0.259 |
| Non-ultra-processed food intake, % of TE (N=1021) | 54.32 (18.45) | 55.22 (25.47) | 53.00 (24.14) | 0.483 |

Data is presented as mean (SD). N-values represent the number of valid cases per dietary risk factor for the total cohort. Values in bold are regarded as statistically significant ( $p \leq 0.05$ ).

### 3.2. Cross-sectional analyses

As presented in Table 3, each 1-gram increment in calcium intake was associated with lower odds of depression in model 1 (OR=0.39; 95% CI: 0.25-0.59), and model 3 (OR=0.48, 95% CI 0.31-0.76), but not in the sociodemographic-adjusted models (models 2 and 4). Each 50-gram increment in wholegrain intake was associated with lower odds of depression in all models: model 1 (OR=0.82, 95% CI 0.67-1.00), model 2 (OR=0.78, 95% CI 0.64-0.95), model 3 (OR=0.82, 95% CI 0.67-1.00) and model 4 (OR=0.78, 95% CI 0.64-0.95). Each 100-gram increment in vegetable intake was associated with lower odds of depression in model 1 (OR=0.74, 95% CI 0.61-0.82), model 2 (OR=0.82, 95% CI 0.67-1.00) and model 3 (OR=0.74, 95% CI 0.61-0.91), but not in model 4. Fibre intake (each 1-gram increment) was associated with lower odds of depression in model 1 (OR=0.97, 95% CI 0.96-0.99) and model 2 (OR=0.98, 95% CI 0.96-1.00), but not in the models adjusted for total energy intake (models 3 and 4).

**Table 3.** Association between dietary intake and the probability of depression.

|  | **PHQ >10** |  |  |
| --- | --- | --- | --- |
| **Model 1** | **OR** | **95% CI** | **p-value** |
| Fruit per 100g | 0.819 | 0.670; 1.000 | 0.057 |
| Vegetables per 100g | 0.740 | 0.606; 0.819 | **<0.001\*** |
| Wholegrains per 50g | 0.818 | 0.669; 1.000 | **0.033** |
| Milk per 10g | 1.000 | 0.990; 1.010 | 0.946 |
| Red meat per 25g | 0.951 | 0.882; 1.000 | 0.064 |
| Processed meat per 17g | 1.017 | 0.983; 1.070 | 0.291 |
| Sugar-sweetened beverages per 50g | 1.000 | 1.000; 1.051 | **0.031** |
| Fibre per 1g | 0.974 | 0.957; 0.992 | **0.005\*** |
| Calcium per 1g | 0.386 | 0.251; 0.592 | **<0.001\*** |
| Omega-3 fatty acids per 1g | 1.000 | 1.000; 1.000 | 0.815 |
| PUFA per % of TE | 0.995 | 0.961; 1.030 | 0.769 |
| UPF per % of TE | 1.004 | 0.996; 1.011 | 0.345 |
| Non-UPF, per % of TE | 0.997 | 0.990; 1.005 | 0.483 |
| **Model 2** | **OR** | **95% CI** | **p-value** |
| Fruit per 100g | 0.905 | 0.740; 1.000 | 0.144 |
| Vegetables per 100g | 0.819 | 0.670; 1.000 | **0.036** |
| Wholegrains per 50g | 0.778 | 0.636; 0.951 | **0.019** |
| Milk per 10g | 1.010 | 1.000; 1.020 | 0.194 |
| Red meat per 25g | 0.951 | 0.905; 1.025 | 0.218 |
| Processed meat per 17g | 1.052 | 1.000; 1.107 | **0.025** |
| Sugar-sweetened beverages per 50g | 1.051 | 1.000; 1.051 | **0.023** |
| Fibre per 1g | 0.977 | 0.958; 0.995 | **0.015** |
| Calcium per 1g | 0.743 | 0.468; 1.180 | 0.209 |
| Omega-3 fatty acids per 1g | 1.000 | 1.000; 1.000 | 0.599 |
| PUFA per % of TE | 0.998 | 0.963; 1.034 | 0.902 |
| UPF per % of TE | 1.005 | 0.998; 1.013 | 0.176 |
| Non-UPF, per % of TE | 0.995 | 0.988; 1.003 | 0.228 |
| **Model 3** | **OR** | **95% CI** | **p-value** |
| Fruit per 100g | 0.905 | 0.740; 1.000 | 0.103 |
| Vegetables per 100g | 0.740 | 0.606; 0.905 | **0.001\*** |
| Wholegrains per 50g | 0.818 | 0.669; 1.000 | **0.029** |
| Milk per 10g | 1.000 | 0.990; 1.010 | 0.560 |
| Red meat per 25g | 0.975 | 0.905; 1.025 | 0.294 |
| Processed meat per 17g | 1.052 | 1.000; 1.107 | 0.056 |
| Sugar-sweetened beverages per 50g | 1.051 | 1.000; 1.051 | **0.004\*** |
| Fibre per 1g | 0.987 | 0.967; 1.007 | 0.167 |
| Calcium per 1g | 0.480 | 0.305; 0.756 | **0.002\*** |
| Omega-3 fatty acids per 1g | 1.000 | 1.000; 1.000 | 0.984 |
| PUFA per % of TE | 0.997 | 0.963; 1.0032 | 0.860 |
| UPF per % of TE | 1.004 | 0.997; 1.012 | 0.296 |
| Non-UPF, per % of TE | 0.997 | 0.990; 1.004 | 0.397 |

| *Model 4* | **OR** | **95% CI** | **p-value** |
| --- | --- | --- | --- |
| Fruit per 100g | 0.905 | 0.740; 1.105 | 0.191 |
| Vegetables per 100g | 0.819 | 0.740; 1.000 | 0.061 |
| Wholegrains per 50g | 0.778 | 0.636; 0.951 | **0.014** |
| Milk per 10g | 1.010 | 1.000; 1.020 | 0.090 |
| Red meat per 25g | 0.975 | 0.905; 1.025 | 0.388 |
| Processed meat per 17g | 1.070 | 1.017; 1.126 | **0.007** |
| Sugar-sweetened beverages per 50g | 1.051 | 0.254; 1.051 | **0.010** |
| Fibre per 1g | 0.982 | 0.961; 1.003 | 0.100 |
| Calcium per 1g | 0.943 | 0.565; 1.574 | 0.822 |
| Omega-3 fatty acids per 1g | 1.000 | 1.000; 1.000 | 0.703 |
| PUFA per % of TE | 0.999 | 0.964; 1.035 | 0.954 |
| UPF per % of TE | 1.006 | 0.998; 1.013 | 0.159 |
| Non-UPF, per % of TE | 0.995 | 0.987; 1.003 | 0.197 |

Model 1 = unadjusted
Model 2 = adjusted for sociodemographic information (age, sex, ethnicity and socio-economic status).
Model 3 = adjusted for total energy intake using Willetts residual method.
Model 4 = adjusted for sociodemographic information and total energy intake using Willetts residual method.
Abbreviations: CI, confidence interval; OR, odds ratio; PUFA, Polyunsaturated fat; UPF, Ultra-processed food intake. Values in bold are regarded as statistically significant ( $p \leq 0.05$ ).
Superscript \* indicates results that remained statistically significant after adjustment for multiple comparisons.

Each 17-gram increment in processed meat consumption was associated with higher odds for depression, but only in the models adjusted for sociodemographic information: model 2 (OR=1.05, 95% CI 1.00-1.11) and model 4 (OR=1.07, 95% CI 1.02-1.13). Intake of sugar-sweetened beverages (each 50-gram increment) was associated with higher odds of depression in all the models: model 1 (OR=1.00, 95% CI 1.00-1.05), model 2 (OR=1.05, 95% CI 1.00-1.05), model 3 (OR=1.05, 95% CI 1.00-1.05) and model 4 (OR=1.05, 95% CI 0.25-1.05).

No associations were observed for fruit, milk, red meat, omega-3 fatty acids, PUFA, UPF or non-UPF in any of the models (Table 3). Additional adjustment for BMI in the sensivitiy model (Model 4 and BMI) did not affect the results in any of the models as shown in Supplementary Table 1. After adjustment for multiple comparisons, associations in model 1 between vegetables, fibre, and calcium and odds for depression remained statistically significant. In model 3, associations with vegetables, sugar-sweetened beverages, and calcium remained significant. In contrast, all associations in models 2 and 4 were no longer statistically significant as shown in Table 3.

### 3.3. Longitudinal analyses

The average follow-up time in this study was 4.96 years. Among the 551 participants included at follow-up, 353 did not meet criteria for depression at either time point. Incident depression was observed in 50 individuals. Remission occurred in 80 participants who met depression criteria at baseline but not at follow-up, while 68 participants met criteria at both assessments. The results of the Poisson regression analyses are shown in Figure 2a-d and Supplementary Table 2. In the total cohort, excluding those who had depression at baseline (n=403), higher sugar-sweetened beverage intake (per 50-gram increment) was associated with a lower risk of incident depression only in the fully adjusted model: model 4 (RR=0.95, 95% CI 0.90-1.00). Milk consumption (per 10-gram increment) was associated with a lower risk of incident depression in model 1 (RR=0.95, 95% CI 0.91-0.99), model 3 (RR=0.94, 95% CI 0.90-0.98) and model 4 (RR=0.95, 95%CI=0.92-0.99), but not in the sociodemographic-adjusted model (model 2). Calcium intake (per 1-gram increment) was association with a lower risk of incident depression in the models that were not adjusted for sociodemographic information: model 1 (RR=0.35, 95% CI 0.13-0.98) and model 3 (RR=0.33, 95% CI 0.11-0.94).

**Figure 2.**
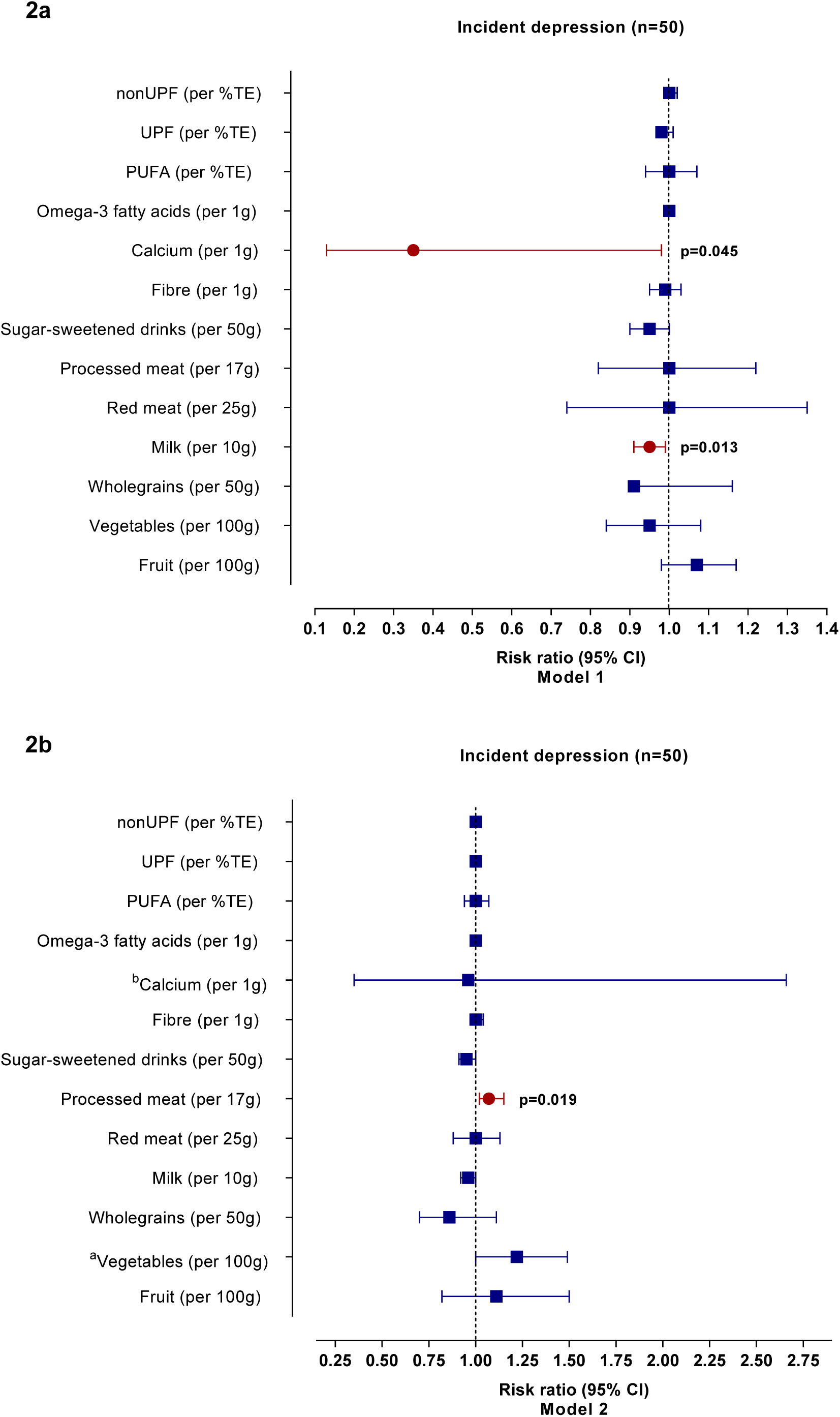
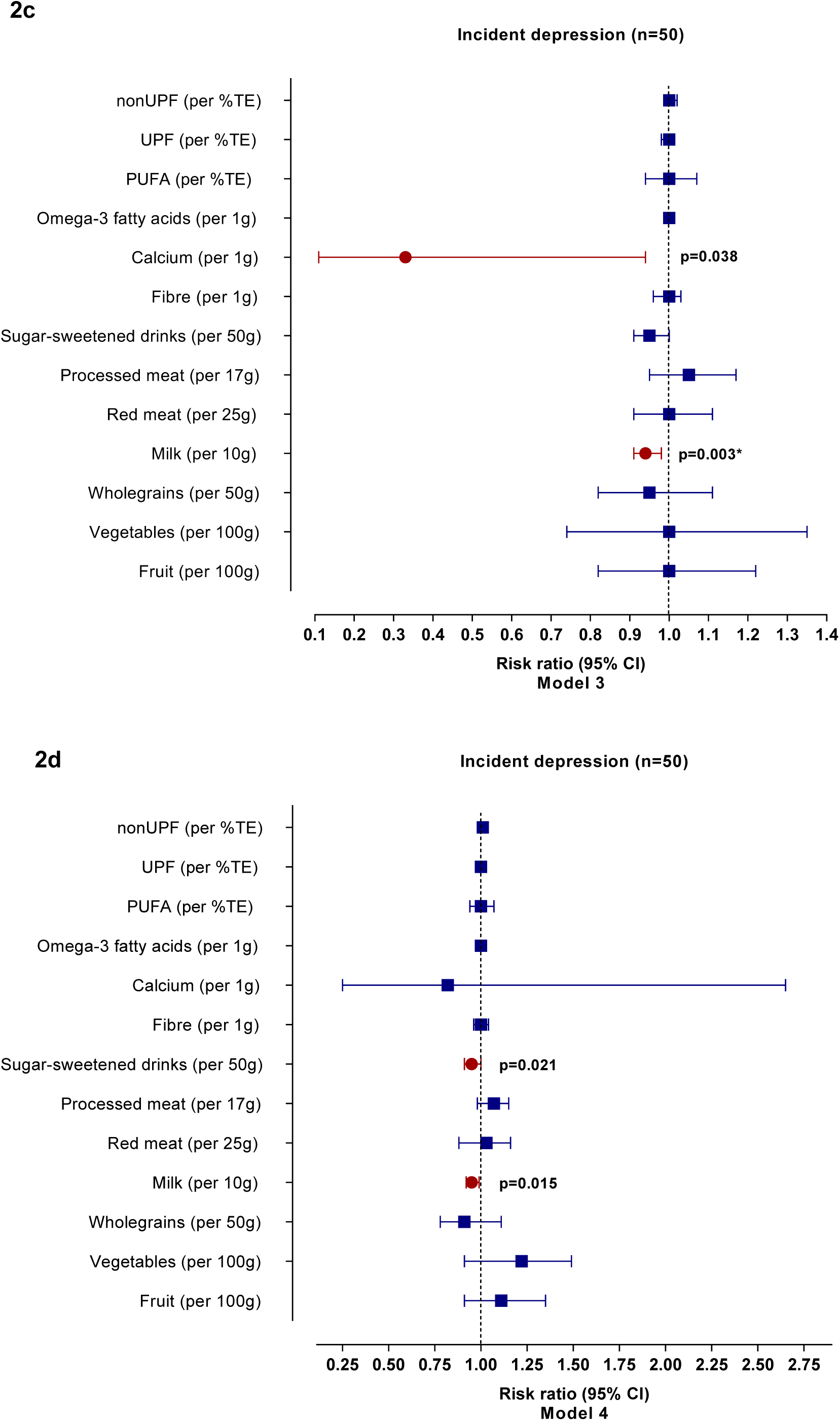
Relative risk of baseline dietary intake predicting incident depression in model 1 (2a), model 2 (2b), model 3 (2c) and model 4 (2d). Abbreviations: g, gram; PHQ, patient health questionnaire; PUFA, poly-unsaturated fatty acids; UPF, ultra-processed foods. Superscript * indicates results that remained statistically significant after adjustment for multiple comparisons.

Processed meat consumption (per 17-gram increment) increased the risk of incident depression in model 2 (RR=1.07, 95% CI 1.02-1.15) but not in the unadjusted model (model 1) or in the models adjusted for TE intake (models 3 and 4).

Consumption of the other dietary exposures in this model, including fibre, omega-3 fatty acids, PUFA and UPF, was not associated with incident depression as shown in Figures 2a-d. Following adjustment for multiple comparisons, all associations in Models 1–2 and 4 lost statistical significance, while only the association with milk in Model 3 remained significant as shown in Figure 2c. Additionally adjusting for BMI in the sensitivity model (adjusting for covariates included in model 4, and BMI) did not affect the outcome of the results in any of the models (Supplementary Table 1).

## 4. Discussion

This study investigated the association between dietary intake -defined according to the GBD study - and symptoms of depression in a cohort of young South African adults. In the cross-sectional analysis, higher intakes of calcium and vegetables were associated with lower odds for depression in the unadjusted models and models adjusted for sociodemographic factors. Higher intakes of fibre were associated with lower odds only in the unadjusted analysis. Higher sugar-sweetened beverage consumption was associated with higher odds for depression, but only after adjustment for sociodemographic factors. In the longitudinal analyses, there was some evidence that higher intake of milk may be associated with a lower risk of incident depression from baseline to follow-up however this only occur when total energy was adjusted for.

### 4.1. Sugar-sweetened beverages as risk factor for depression

Cross-sectionally, sugar-sweetened beverage consumption was associated with higher odds for depression in the models adjusted for TE intake consistent with previous findings [26–28]. Increased sugar-sweetened beverage consumption has been linked to reduced dopaminergic brain responses in the posterior midbrain and dorsolateral prefrontal cortex of young adults [29], regions suspected to be implicated in the pathophysiology of depression. These findings appear to be driven by the increased intake of sugar content rather than total caloric-intake [29], which aligns with our finding that it only became significant after adjusting for TE intake. High–sugar diets may contribute to depression through multiple biological pathways, including disrupted insulin signalling, hypothalamic-pituitary-adrenal (HPA)-axis dysregulation, systemic inflammation, impaired neuroplasticity and neurotransmitter function, and alterations in the gut–brain axis. Collectively, these effects can interfere with brain regions and signalling systems critical for mood regulation [30].

In contrast to the cross-sectional findings, longitudinal analyses suggested that sugar-sweetened beverage consumption at baseline was associated with a lower risk of incident depression over time which is inconconsistent with the the vast majority of the literature and lacks biological plausability. It is therefore likely to be either a spurios finding owing to statistical chance, small sample size at follow-up, or methodological limitations - particularly given that the association was observed only in the model adjusted for both TE intake and sociodemographic factors, and did not remain significant after correction for multiple comparisons. For example, we did not capture changes in dietary risk factors over time to know whether SSB consumption reduced over the study period which may have helped explain this protective finding. It is plausible that population level reductions in SSB occurred - notably, sugar-sweetened beverage intake decreased by approximately 33% between 2018 and 2023 [31], a trend also observed in South African children living in the same geographical area as the young adults in our cohort [32]. Such a decline would be consistent with existing evidence suggesting that reductions in sugar-sweetened beverage intake may have beneficial effects on mental health outcomes. This may particularly be true for young adults as they were reported to consume more sugar-sweetened beverages than other age groups [33]. Furthermore, young adults with depression in our cohort reported a higher consumption of sugar-sweetened beverage consumptions when compared to those without depression.

The discrepancy in the direction of associations observed between the cross-sectional and longitudinal analyses may also reflect reverse causality in the cross-sectional findings, as causal inferences cannot be drawn from cross-sectional data alone. In our cohort, individuals with depression reported higher consumption of sugar-sweetened beverages, which may indicate not only a potential contribution of these beverages to depression risk, but also that individuals with depressive symptoms consumed them more frequently as a potential coping mechanism.

Indeed, previous findings among young adults have shown that the cross-sectional association between depressive symptoms and sugar consumption is mediated by emotional dysregulation, including emotional eating and food cravings [34].

### 4.2. The association between processed meat consumption and depression

In the sociodemographic-adjusted cross-sectional and longitudinal models, processed meat consumption was positively associated with depression, in line with previous research [35]. Processed meat, usually defined as products made of red meat that has been cured, salted or smoked to extend shelf-life, is generally high in saturated fat, sodium, nitrites/nitrates, and pro-oxidant compounds such as advanced glycation end-products (AGEs) [36]. Such a profile may promote inflammation and oxidative stress thereby increasing the risk of depression. In contrast to a study that found a higher intake of processed meat to increase the risk of developing late-onset depression in older adults over a 12-year period [37], this association was not observed in the unadjusted or energy-adjusted-only models, suggesting that sociodemographic factors may confound the relationship. Indeed, previous studies in South Africa have shown that processed meat consumption is higher among men, Black individuals, those with higher SES, and young adults [38]. Furthermore, the findings with processed meat consumption were no longer significant after multiple comparison corrections and therefore must be interpreted with caution.

### 4.3. Calcium intake and milk consumption as potential protective factors for depression

Several micronutrient deficiencies contribute to the pathophysiology of depression. Consistent with our cross-sectional and longitudinal findings, several studies have reported an inverse association between dietary calcium intake and the prevalence of depressive symptoms [39–41]. Calcium is essential for neurotransmission through its involvement in various types of calcium channels that regulate presynaptic neurotransmitter release, postsynaptic signalling, and synaptic plasticity [42]. Disruptions in long-term synaptic plasticity, particularly in the prefrontal cortex and hippocampus, have been implicated in depression [43]. Inadequate calcium intake may therefore impair synaptic transmission and plasticity, contributing to depressive pathology. Calcium intake was lower in those with depression compared to individuals without depression at baseline, underscoring the potential importance of calcium deficiencies. Sociodemographic context appears particularly important, as these findings were not observed in the models adjusted for sociodemographic factors, indicating that the associations may reflect influences of specific sexes, races, socioeconomic groups, or interactions among these factors. In our study, participants with depression were predominantly Black and of low SES. Furthermore, previous African-PREDICT findings showed that Black individuals had lower calcium intakes than White individuals, although neither group met the estimated average calcium requirements, with more than 50% of the entire cohort across all SES groups not meeting these levels [44].

Similarly, milk consumption was associated with a lower risk of incident depression in the longitudinal analyses. Although the specific mechanisms underlying the inverse findings remains unclear as if reflected by the contradicting results reported on the relationship between depression and milk intake in the literature [45], the nutritional composition of milk may be relevant. Milk provides a substantial source of dietary calcium, supporting both calcium intake and levels. The chief protein in mammalian milk, casein, supports calcium absorption and provides amino acids important for neurotransmitter synthesis, and has been shown, in combination with GABA, to reduce depressive-like behaviour in animal models [46, 47]. Fat content may also be relevant, as low-fat varieties, such as semi-skimmed and skimmed milk, are associated with a lower risk of depression, whereas whole milk may increase the risk [48, 49]. However, we did not observe these findings in our cross-sectional analyses, suggesting that the effects of inadequate milk intake on depressive symptoms may only become noticeable after a longer period of time, with limited measurable effects detectable in the short term. Indeed, young adulthood may represent a key developmental period during which the effects of inadequate milk intake on depression are not immediately observable but emerges over time. Consistent with this, no differences in milk consumption were observed between individuals with and without depression at baseline, which may explain why associations were only observed in the longitudinal analyses. Longitudinal assessment of dietary changes would therefore further clarify these inverse associations.

### 4.4. The inverse associations between wholegrain, fibre, and vegetable consumption with depression

We observed inverse cross-sectional associations between depression and intake of wholegrains, dietary fibre, and vegetables, partially supporting previous evidence that higher consumption of these foods may be protective against depressive symptoms [50–53]. However, none of these associations were observed in the longitudinal analyses, and most were no longer significant after adjustment for TE intake or correction for multiple comparisons. It is also possible that the cross-sectional associations may reflect reverse causality as individuals with depression exhibited lower consumption of wholegrains, fibre, and vegetables when compared to those without, or residual confounding rather than a sustained effect over time. Overall, these findings highlight the need for cautious interpretation and underscore the challenges of disentangling dietary influences on depression in young adults, where evidence remains inconsistent.

### 4.5. Strengths and limitations

This study offers valuable insights into the relationship between dietary intake and depression, laying a foundation for future longitudinal research to assess whether dietary interventions could improve depressive symptoms. However, certain limitations must be acknowledged. The African-PREDICT study included a screened cohort of apparently healthy young adults without overt cardiovascular disease, all residing within a specific geographical area. As a result, the findings may not be generalizable to the broader South African population. While validated tools were used to assess the presence and severity of depression, the use of a structured clinical interview would have strengthened the reliability of the findings by capturing symptom duration beyond the preceding two weeks.

### 4.6. Conclusion

Milk consumption at baseline was associated with a lower risk of incident depression at follow-up. In addition, depression associated positively with sugar-sweetened beverages and inversely with vegetables, fibre and calcium in cross-sectional analyses. Establishing healthy dietary habits early in life by ensuring adequate daily milk, calcium, fibre and vegetable intake may have potential to help reduce the risk of depression young adults. These findings highlight the importance of incorporating dietary considerations into mental health prevention strategies.

## Data Availability

All data produced in the present study are available upon reasonable request to the corresponding author.

## Acknowledgements

The authors would like to acknowledge and thank Mrs D Kruger from the Pure and Applied Analytics, School of Mathematical and Statistical Sciences, North-West University, Potchefstroom, South Africa, for conducting the initial, preliminary statistical analysis. The research funded in this manuscript is part of an ongoing research project financially supported by the South African Medical Research Council (SAMRC) with funds from National Treasury under its Economic Competitiveness and Support Package and SAMRC Extramural Research Unit funding (SAMRC-RFA-EMU-10-2020); the South African Research Chairs Initiative (SARChI) of the Department of Science and Technology and National Research Foundation (NRF) of South Africa (GUN 86895); SAMRC with funds received from the South African National Department of Health, GlaxoSmithKline R&D (Africa Non-Communicable Disease Open Lab grant), the UK Medical Research Council and with funds from the UK Government’s Newton Fund; as well as corporate social investment grants from Pfizer (South Africa), Boehringer-Ingelheim (South Africa), Novartis (South Africa), the Medi Clinic Hospital Group (South Africa) and in kind contributions of Roche Diagnostics (South Africa). Any opinion, findings, and conclusions or recommendations expressed in this material are those of the authors; therefore, the NRF does not accept any liability in this regard. The GLAD project is funded by a National Health and Medical Research Council Emerging Leader 2 Fellowship (2009295 [AO]).

## 5. Author contributions

EJvV: Data analysis and interpretation, Writing – original draft, review and editing; AON: Conceptualization, Methodology - developing the methods for the GLAD project, Project administration – management and coordination of the GLAD project, Resources - provision of study materials, including the data analysis code and materials of the GLAD project, Writing – review and editing; DNA: Conceptualization, Methodology - developing the methods for the GLAD project, Project administration – management and coordination of the GLAD project, Resources - provision of study materials, including the data analysis code and materials of the GLAD project, Writing – review and editing; MML: Conceptualization, Methodology - developing the methods for the GLAD project, Project administration – management and coordination of the GLAD project, Resources - provision of study materials, including the data analysis code and materials of the GLAD project, Writing – review and editing; RO: Conceptualization, Methodology - developing the methods for the GLAD project, Project administration – management and coordination of the GLAD project, Resources - provision of study materials, including the data analysis code and materials of the GLAD project, Writing – review and editing; MP: Writing – review and editing; TvZ: Sample and data analysis, writing – review and editing.

All authors read and approved the final manuscript.

---

**Supplementary Table 1.** Sensitivity analyses of associations between dietary intake and depression after additional adjustment for BMI

| **n=306** |  |  |  |
| --- | --- | --- | --- |
| **Probability for depression** | **OR** | **95% CI** | **p-value** |
| Fruit per 100g | 0.905 | 0.740; 1.105 | 0.187 |
| Vegetables per 100g | 0.819 | 0.740; 1.000 | 0.062 |
| Wholegrains per 50g | 0.778 | 0.605; 0.951 | **0.014** |
| Milk per 10g | 1.010 | 1.000; 1.020 | 0.085 |
| Red meat per 25g | 0.975 | 0.905; 1.025 | 0.281 |
| Processed meat per 17g | 1.070 | 1.017; 1.126 | **0.007** |
| Sugar-sweetened beverages per 50g | 1.051 | 1.000; 1.051 | **0.011** |
| Fibre per 1g | 0.984 | 0.962; 1.005 | 0.137 |
| Calcium per 1g | 0.961 | 0.576; 1.605 | 0.880 |
| Omega-3 fatty acids per 1g | 1.000 | 1.000; 1.000 | 0.701 |
| PUFA per % of TE | 0.998 | 0.963; 1.034 | 0.925 |
| UPF per % of TE | 1.006 | 0.998; 1.014 | 0.148 |
| Non-UPF, per % of TE | 0.995 | 0.987; 1.002 | 0.182 |
| **n=50** |  |  |  |
| **Relative risk for incident depression** | **RR** | **95% CI** | **p-value** |
| Fruit per 100g | 1.105 | 0.905; 1.349 | 0.305 |
| Vegetables per 1g | 1.221 | 0.905; 1.491 | 0.123 |
| Wholegrains per 50g | 0.905 | 0.740; 1.162 | 0.438 |
| Milk per 10g | 0.961 | 0.923; 0.990 | **0.018** |
| Red meat per 25g | 1.000 | 0.891; 1.133 | 0.915 |
| Processed meat per 17g | 1.070 | 0.983; 1.145 | 0.134 |
| Sugar-sweetened beverages per 50g | 0.951 | 0.905; 1.000 | **0.031** |
| Fibre per 1g | 1.001 | 0.960; 1.043 | 0.979 |
| Calcium per 1g | 0.869 | 0.267; 2.826 | 0.816 |
| Omega-3 fatty acids per 1mg | 1.000 | 1.000; 1.000 | 0.461 |
| PUFA per % of TE | 1.000 | 0.939; 1.066 | 0.996 |
| UPF per % of TE | 0.996 | 0.984; 1.008 | 0.510 |
| Non-UPF, per % of TE | 1.004 | 0.992; 1.017 | 0.497 |

Adjusted for body mass index in addition to sociodemographic information and total energy intake using Willetts residual method.
Abbreviations: CI, confidence interval; OR, odds ratio; PUFA, Polyunsaturated fat; UPF, Ultra-processed food intake. Values in bold are regarded as statistically significant ( $p \leq 0.05$ ).
Superscript \* indicates results that remained statistically significant after adjustment for multiple comparisons.

**Supplementary Table 2.**
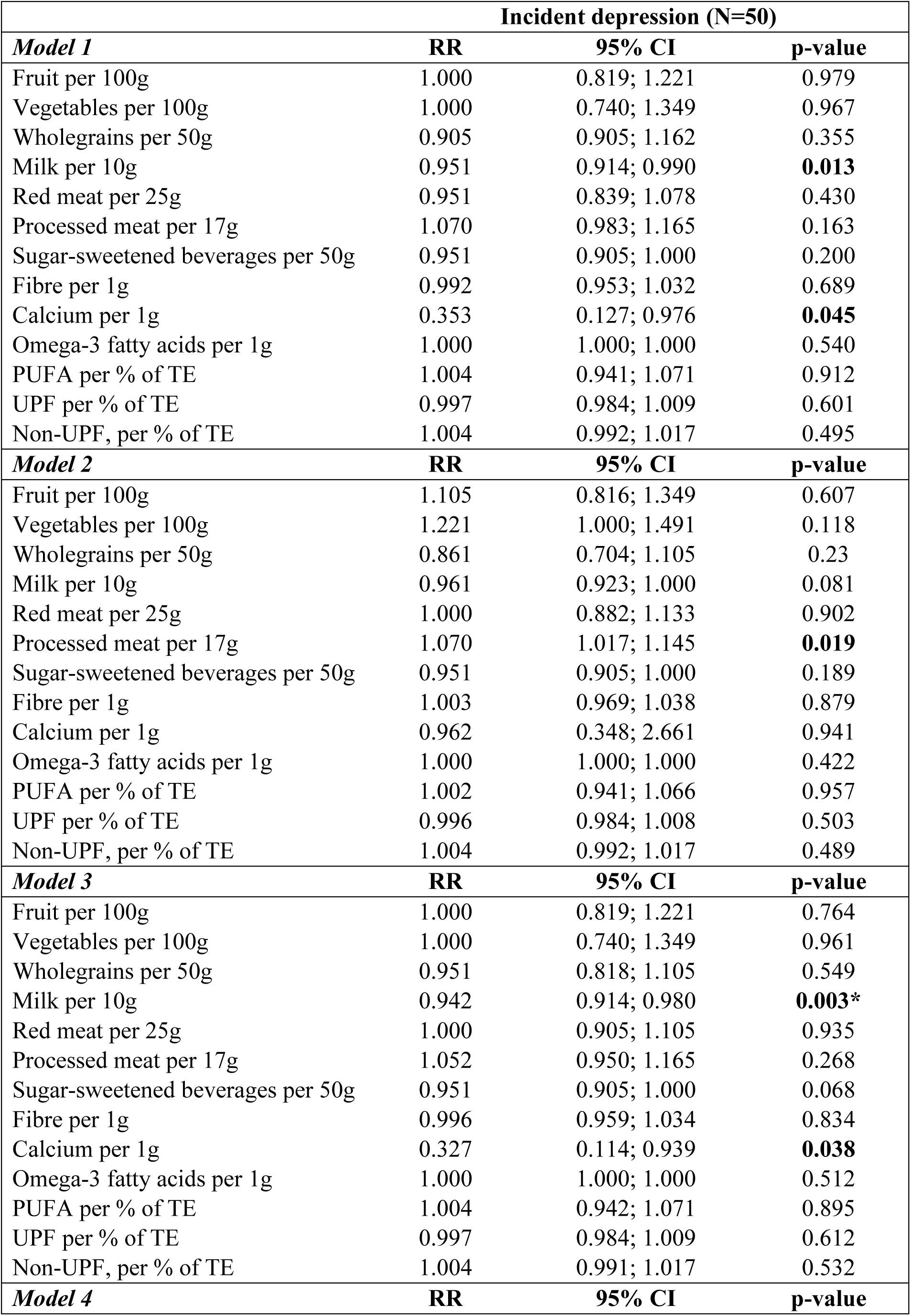
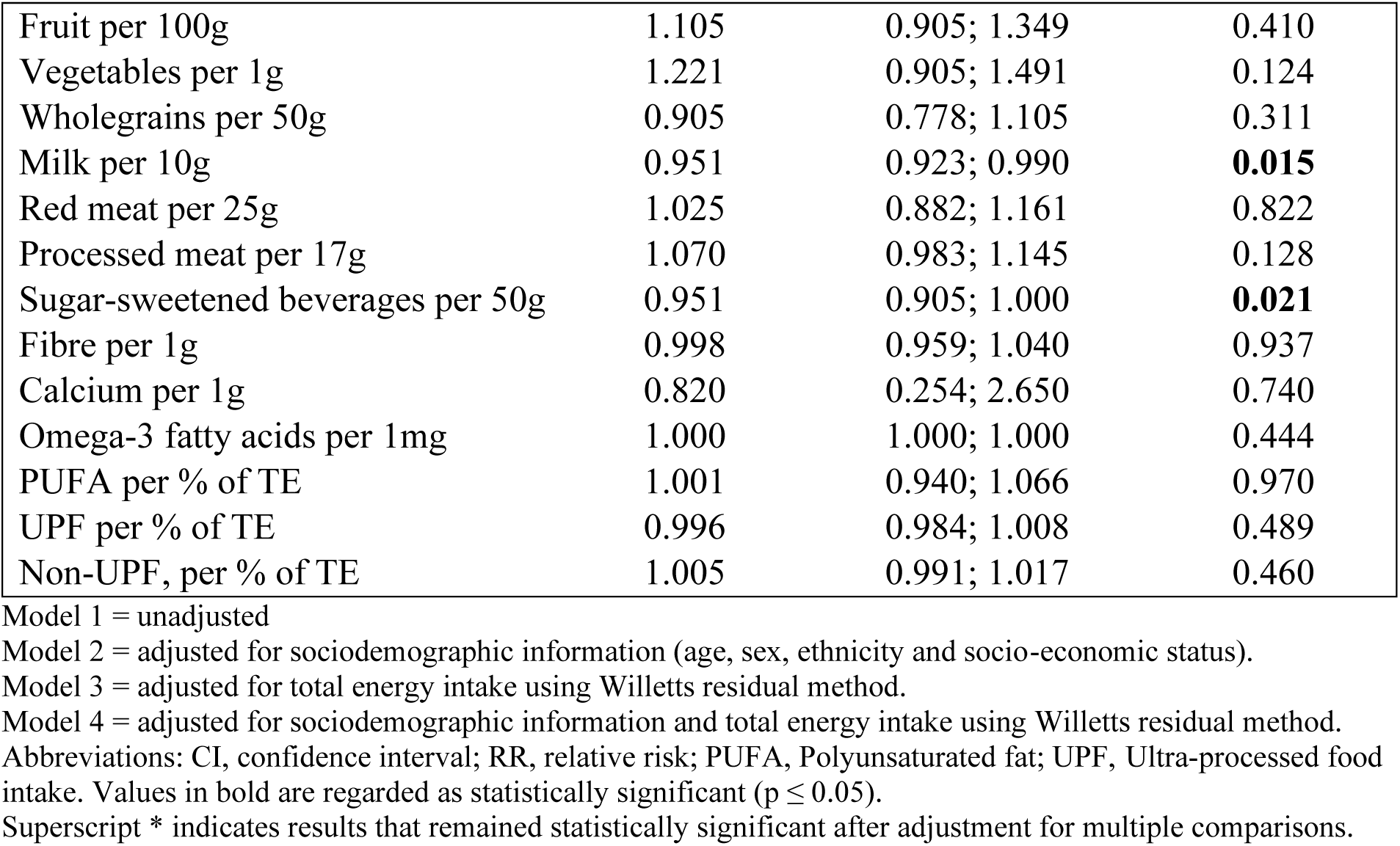
Association between dietary intake and the relative risk of incident depression between baseline and follow-up

